# Real-World Outcomes of a Telephone-Based Virtual Cognitive Rehabilitation Therapy Program: A Retrospective Cohort Analysis

**DOI:** 10.64898/2026.07.10.26357703

**Authors:** Jennifer Flexman, Jessie Ng, Erik Risinger, Corinna Serviente, Michael Busa

## Abstract

**Background:** Cognitive rehabilitation (CR) is an established behavioral intervention that improves daily functioning for individuals with mild cognitive impairment (MCI) and early-stage dementia. Traditional models of in-person delivery limit access, particularly for individuals living in rural areas. This study evaluated the effectiveness of a novel fully remote, telephone-based CR model combining speech-language pathologist (SLP)-led sessions and automated voice-agent based cognitive exercise sessions.

**Methods:** We conducted a retrospective observational analysis of 141 older adults referred for CR who completed treatment to discharge (58% female; mean age 71.2 ± 10.8 years; MCI diagnosis rate 61.7%, dementia diagnosis rate 29.1%; Montreal Cognitive Assessment mean score 21.1 ± 4.0). Changes in four outcome measures from initiation of treatment to discharge were evaluated for statistical significance: quality of life and three therapist-rated Functional Communication Measures (FCMs) of overall cognition, language expression, and language comprehension. Changes were compared to FCM averages from the American Speech-Language-Hearing Association National Outcomes Measurement System (ASHA NOMS). Models were developed to predict changes in outcome measures based on patient demographics, clinical status, program engagement and treating therapist.

**Results:** All four outcomes improved significantly over the course of treatment (p<0.05), with medium to very large effect sizes. Mean changes in FCM outcomes exceeded comparable ASHA NOMS benchmarks for in-person outpatient care. A majority of patients saw an improvement in each clinical outcome measure. Models with meaningful predictive power were identified for changes in overall cognition and language expression, and a statistically significant but weak model was identified for quality of life. No predictive model was identified for spoken language comprehension. Baseline cognitive function was the most influential and negatively correlated predictor of an improvement in overall cognitive abilities and language expression. Baseline quality of life was the dominant and negatively correlated predictor of improvement in quality of life.

**Conclusions:** Telephone-based virtual CR led by SLPs with automated exercises delivered by a voice agent produced clinically meaningful gains, with functional gains comparable to external benchmarks for in-person clinical practice. These results support the use of virtual CR within post-diagnostic care for older adults experiencing cognitive impairment, with greater benefit observed among patients with greater baseline impairment. These results support the expanded use of virtual CR particularly for patients in rural or underserved communities.

## Introduction

Cognitive impairment affects 32% of adults over 65 years old, impacting approximately 250 million people globally^1,2^. Cognitive rehabilitation (CR) is an established behavioral intervention led by skilled therapists to improve cognitive function for individuals experiencing mild cognitive impairment (MCI) and the early stages of dementia. CR works in part through helping individuals and their care partners or family members to overcome practical barriers associated with cognitive difficulties in areas such as memory, task execution, concentration, and communication.

Using a personalized approach focused on the most challenging areas of daily life identified by the individual, CR focuses on enabling individuals to make the most of their remaining abilities and become more independent in everyday life. Speech-language pathologists (SLPs) deliver CR, teaching compensatory strategies and providing targeted cognitive exercises as part of a personalized care plan focused on the individual’s goals and deficits. Therapists help individuals to translate learning and behavioral change into daily life for sustained benefit.

The evidence base shows that individuals living with mild-to-moderate dementia can improve their ability to manage activities of daily life targeted through cognitive rehabilitation^3^. Better cognitive function has been associated with an improvement in the ability to perform activities of daily living in patients with mild-to-moderate Alzheimer’s disease^4,5^. Individualized CR has been shown to have a meaningful impact on individuals and their families coping with mild-to-moderate dementia, preserving everyday functioning, delaying institutional care by six months, and reducing caregiver burden^6–8^. A recent study showed that a combination of cognitive training, lifestyle modules and coaching resulted in sustained improvement in cognitive function, connectedness and emotional balance even in healthy adults^9^.

Despite the growing body of evidence showing the benefits of incorporating CR into dementia care, access to this type of therapy remains limited due to a lack of awareness and availability of skilled practitioners^10,11^. Access gaps are particularly acute in low-resource and remote settings, where comprehensive neurological care may be, at best, hours away. The use of telehealth delivery can bring CR into the home and address gaps in care created by geography, workforce shortages and mobility challenges. In particular, telephone-based therapy has the potential to address technology literacy and broadband barriers associated with video-based delivery and aligns with a preference of older adults more broadly for audio-based telehealth^12^.

The delivery of CR entirely over the telephone has the potential to expand access to patients without access to in-person care, particularly in rural areas, and extend the reach of therapists through the use of automated audio-based cognitive exercises. In this model of virtual CR, patients receive interactive cognitive exercises selected by their therapist and delivered by a voice agent between live sessions with a therapist, all carried out over a basic telephone. Automated cognitive exercises are clinically-designed, interactive question-and-response activities that stimulate areas of cognitive deficit, teach compensatory strategies and allow patients to build confidence in communication in a simulated environment. While there are clear access and convenience benefits for virtual CR, the effectiveness of this model has yet to be evaluated.

This study aims to quantitatively assess changes in clinical outcomes using real-world data, based on the recent deployment of a virtual CR program supporting older adults under Medicare reimbursement in the United States. The purpose of the study was to determine if telephone-based virtual CR combined with automated voice-based activities resulted in significant improvements in patient-reported quality of life and therapist-assessed cognitive function in a patient sample of individuals living with early-stage cognitive decline. As an exploratory analysis, this study also evaluated if factors such as demographics, clinical status, program engagement and treating therapists were reliable predictors of changes in clinical outcomes.

## Methods and Study Design

### Setting and Participants

A retrospective analysis was conducted with a cohort of patients who completed a telephone-based program for virtual cognitive rehabilitation in the United States led by licensed SLPs, upon referral from a physician or nurse practitioner. Cohort patients were initially screened for functional hearing level over the telephone, presence of cognitive complaints, willingness to participate and fluency in English. At the start of therapy, a comprehensive evaluation was completed by the treating SLP to determine baseline cognitive functional status, deficits by cognitive domain, and identify goals to inform treatment planning by the therapist. Care partners such as family members were invited to optionally participate through attending sessions with the patient and assisting the patient during the program.

The therapy care plan consisted of two 20-30 minute sessions over the telephone with the SLP per week for up to 24 weeks. Patients were also asked to complete at least three sessions of cognitive exercises each week, which were delivered to the patient through an automated system over the telephone. These sessions were 15-20 minutes each and composed of three cognitive exercises selected by the treating SLP based on the patient’s goals and to address the patient’s cognitive challenges, such as for short term memory and executive functioning. Cognitive exercises also incorporated therapist-derived content to teach cognitive strategies for everyday life. Automated calls were recorded, transcribed, and reviewed by the SLP as part of treatment monitoring through a web-based software portal. Cognitive exercises were updated for the patient on a weekly basis by the SLP based on patient feedback and performance. Each month, the treating SLP recertified the patient to continue based on an assessment of continued progress towards goal areas.

### Ethics Approval

The Institutional Review Board at the University of Massachusetts Amherst granted a human subjects research waiver prior to conducting this retrospective analysis as the study did not involve intervention or interaction with individuals and did not involve identifiable private information. The study was conducted in accordance with the Declaration of Helsinki. To ensure the privacy and confidentiality of participants, all data were anonymized before analysis, and patient consent was therefore waived. No financial compensation was provided to individuals for their involvement in this study.

### Data Collection and Outcome Measures

A retrospective observational study was designed to analyze four outcome measures collected during routine virtual CR care at the time of initial evaluation (PRE) and at discharge (POST) to evaluate changes (Δ) over the course of treatment, shown in Table 1. Functional communication measures were assessed only if appropriate for cognitive domains targeted by the care plan. As a result, sample sizes varied across measures due to differential rates of assessment completion.

**Table 1.** Overview of outcome measures. FCM = functional communication measure; SLP = speech-language pathologist; SD = standard deviation.

| Name | Measure | Reporter | Instrument |
| --- | --- | --- | --- |
| **Neuro-QoL T-Score** | T-score for self-reported cognitive functioning, normed to a population mean±SD of 50±10. | Patient | Quality of Life (QoL) in Neurological Disorders Item Bank v2.0 - Cognition Function, Short Form<sup>13</sup> |
| **FCM-Cognition** | Functional assessment of overall cognitive abilities in daily life rated on four factors and converted to a 0-100 score | Trained SLP | Functional Communication Measure (FCM) for Cognition for ages 6+<sup>14</sup> |
| **FCM-Language Comprehension** | Functional assessment of ability to understand language in daily life rated on six factors and converted to a 0-100 score | Trained SLP | FCM for Spoken Language Comprehension for ages 6+<sup>14</sup> |
| **FCM-Expressive Language** | Functional assessment of use of verbal skills in daily life rated on seven factors and converted to a 0-100 score | Trained SLP | FCM for Spoken Language Expression for ages 6+<sup>14</sup> |

### Analysis of Pre-post Changes

The magnitude and significance of change, and 95% confidence interval, were analyzed for all four outcome measures from baseline (PRE) to discharge (POST). Statistical analysis of pre-post score changes was conducted using SciPy, NumPy and pandas. Statistical significance of score changes from PRE to POST was assessed using a paired sample t-test (Neuro-QoL) or a Wilcoxon signed-rank test (FCMs) based on the measurement level of each instrument (continuous T-score versus ordinal FCM ratings) with a significance level of *p*<0.05. Effect size magnitudes (Cohen’s *d*) were interpreted as very small (0.01), small (0.2), medium (0.5), large (0.8), very large (1.2), and huge (2.0)^15^.

Changes in FCM outcome measures observed in this study were compared to benchmark data in the American Speech-Language-Hearing Association National Outcomes Measurement System (ASHA NOMS) database, a voluntary SLP registry available to US clinicians holding ASHA membership to record anonymous FCM data at admission and discharge^16^. Benchmarks for changes in FCMs were extracted for patients in outpatient in-person settings for cognitive communication disorders, ages 30-89 years old.

### Outcome Predictors

Thirty-five candidate features were derived from intake records and program engagement logs across four domains: (1) *demographics* – age, sex, level of education; (2) *clinical characteristics* – diagnoses, hearing impairment status, living environment, Global Deterioration Scale (GDS) level, Montreal Cognitive Assessment (MoCA)-BLIND version 8.1 score (converted to 30-point scale^17^), Ross Information Processing Assessment - Geriatric, Second Edition (RIPA-G:2) raw score excluding the spatial orientation subtest, and baseline Neuro-QoL (raw and T-score) and FCM-Cognition scores; (3) *program engagement* – total automated therapy call count, average weekly automated therapy calls, and number of weeks enrolled; and (4) *therapist assignment* – a nominal code identifying one of five treating speech-language pathologists. Seven patients had no treating therapist recorded and were therefore excluded from modeling by listwise deletion. For the same reason, baseline FCM-Expressive Language and FCM-Language Comprehension scores, recorded for only 85 and 54 of the 141 patients, were not included as predictors: retaining both would have reduced every target to 30 or fewer complete cases, fewer than the number of candidate predictors. Several characteristics entered in more than one encoding: living environment as six indicator variables plus an ordinal grouping; GDS as its level plus two binary groupings (level 4 or 5 versus 2 or 3, and level 3 or higher versus 2); and MoCA as its score plus a four-level ordinal severity grouping.

### Comparative Modeling of Outcome Measure Prediction

Models were developed to predict changes in each of the outcome measures listed in Table 1 based on predictors. Given the limited sample size, a structured model comparison framework was used emphasizing methodological rigor and controlling for overfitting. The feature set included continuous, ordinal, nominal, and binary variables. The treating-therapist code was one-hot encoded with one category omitted as a reference to prevent multicollinearity, and living environment was represented by six indicator variables (one of which, assisted living, was unpopulated in this cohort); binary variables were encoded as 0/1; ordinal and continuous variables were retained in numeric form. Standardization (StandardScaler) was performed independently within each cross-validation fold. No imputation was conducted for outcome or continuous predictor values; participants with incomplete outcome or predictor data for a given target were excluded on a per-target basis, yielding analytic sample sizes ranging from 48 to 128 across four outcome measures. Patients with unknown hearing status (n = 8) were coded as having no hearing impairment.

All models were estimated within a comparative modeling framework on identical 5-fold × 10 repeat cross-validated (CV) folds within each target. Five model classes were evaluated per outcome: Gradient Boosting, Ridge regression, Lasso, Elastic Net, and Bayesian Ridge, alongside a single-feature baseline using the strongest bivariate predictor. Feature selection for linear models was conducted via nested CV combined with recursive feature elimination (RFE), sweeping candidate subset sizes of 3, 5, 7, and 10 features; the configuration maximizing mean CV *R*^*2*^ was retained.

Primary performance metrics were mean CV *R*^*2*^ and root mean squared error (RMSE). Statistical significance was assessed via permutation testing (1,000 permutations of the outcome vector; reported p-values are one-tailed) with significance at *p*<0.05. Ninety-five percent confidence intervals (CIs) were constructed via bootstrap resampling of CV fold scores; CIs crossing zero are considered consistent with chance. An interval lying entirely below zero indicates that a model reliably underperforms the sample mean across folds, which is a separate question from whether it outperforms permuted outcomes. Where ordinary least squares (OLS) assumptions were satisfied (Shapiro-Wilk normality and Levene homoscedasticity tests on residuals from a univariate OLS fit on the continuous predictor most strongly correlated with the outcome), the Akaike Information Criterion corrected for small samples (AICc) was additionally reported for the linear model family.

Cross-validated *R*^2^ was the primary model selection criterion across all model families, and was interpreted as follows: <0 (no predictive value), <0.05 (negligible), <0.1 (weak), and ≥0.1 (modest/meaningful).

## Results

### Sample Selection and Demographic Characteristics

The study population included 141 individuals who completed treatment through to discharge (n=141, 42% male and 58% female; mean age of 71.2±10.8 years). Of 220 patients considered for analysis, patients that did not complete at least four weeks of the program (39/220, 17.7%) or for whom discharge assessment data were not available (40/220, 18.2%) were not included in the sample. See Table 2 for full study population characteristics. Correlation of study population characteristics is shown in Supplemental Data - Figure A5.

**Table 2.** Characteristics of study population. (N = 141). SD = standard deviation; MCI = mild cognitive impairment; FCM = functional communication measure.

| Characteristic | Representation in Dataset |
| --- | --- |
| **Age**, mean ± SD (range) | $71.2 \pm 10.8$ (37.0–88.0) |
| **Sex**, n (%) | Female: 82 (58.2%); Male: 59 (41.8%) |
| **Has care partner such as a family member**, n (%) | 44 (31.2%) |
| **Level of education**, n (%) | Did not finish high school: 6 (4.3%)<br>Finished high school: 24 (17.0%)<br>High school with 1–3 years post-secondary: 52 (36.9%)<br>High school with 4+ years post-secondary: 53 (37.6%)<br>Unknown: 6 (4.3%) |
| **Current living environment**, n (%) | Independently with family: 62 (44.0%)<br>At home with family support: 42 (29.8%)<br>Alone and independently: 31 (22.0%)<br>At home with paid support: 3 (2.1%)<br>Supportive care community: 3 (2.1%) |
| **Diagnoses** (ICD-10 coding), n (%)<br>*Note: diagnoses are not exclusive, but patients cannot have a diagnosis of MCI and dementia.* | *Cognitive diagnoses:*<br>MCI: 87 (61.7%)<br>Dementia (excluding MCI): 41 (29.1%)<br>Unknown diagnosis: 1 (0.7%)<br>Other diagnosis: 12 (8.5%)<br>*Secondary diagnoses:*<br>Cerebrovascular accident (CVA) / stroke: 8 (5.7%)<br>Traumatic brain injury (TBI) / post-concussive: 9 (6.4%)<br>Multiple sclerosis (MS): 3 (2.1%)<br>Parkinson's disease (PD): 11 (7.8%) |
| **Hearing impairment**, n (%) | Yes: 53 (37.6%); Unknown: 8 (5.7%) |
| **Montreal Cognitive Assessment (MoCA) BLIND**, raw score converted to 30-point scale, mean ± SD (range), N=135 | 21.1 ± 4.0 (8.0–28.0) |
| **Ross Information Processing Assessment - Geriatric, Second Edition (RIPA-G:2)** raw score excluding the spatial orientation subtest, mean ± SD (range) | 225.2 ± 33.6 (101.0–291.0) |
| **Global Deterioration Scale (GDS)** level, n (%)<br>*Related parameters tested:*<br>'GDS Grouping 1' classes: 0 = GDS level of 2 or 3; 1 = GDS level of 4 or 5<br>'GDS Grouping 2' classes: 0 = GDS level of 2; 1 = GDS level of 3, 4 or 5 | Level 2: 11 (7.8%)<br>Level 3: 102 (72.3%)<br>Level 4: 27 (19.1%)<br>Level 5: 1 (0.7%) |
| **Program duration** in weeks, mean ± SD (range) | 9.6 ± 4.4 (4.0–24.0) |
| **Total automated therapy calls**, total calls completed during treatment > 5 min in duration, mean ± SD (range), N=132 | 25 ± 18 (3–98) |
| **Average weekly automated calls**, average calls > 5 min in duration completed per week in treatment, mean ± SD, N=132 | 2.7 ± 0.8 |
| **Neuro-QoL T-Score (PRE)**, mean ± SD (range), N=141 | 38.4 ± 6.3 (17.3–56.3) |
| **Neuro-QoL T-Score, change (POST-PRE)**, mean ± SD (range), N=116 | 4.0 ± 5.6 (-10.1 to 21.5) |
| **FCM-Cognition (PRE)**, mean ± SD (range), N=137 | 65.3 ± 24.8 (0.0–100.0) |
| **FCM-Cognition, change (POST-PRE),**<br>mean ± SD (range), *N*=134 | 18.2 ± 18.6 (-13.0 to 81.0) |
| **FCM-Expressive Language (PRE),**<br>mean ± SD (range), *N*=85 | 71.9 ± 16.3 (36.0–96.0) |
| **FCM-Expressive Language, change (POST-PRE),** mean ± SD (range),<br>*N*=83 | 13.8 ± 14.1 (-36.0 to 50.0) |
| **FCM-Language Comprehension (PRE),** mean ± SD (range), *N*=54 | 62.1 ± 21.5 (8.0–96.0) |
| **FCM-Language Comprehension, change (POST-PRE),** mean ± SD (range), *N*=54 | 18.8 ± 14.4 (-21.0 to 50.0) |
| **Therapist code** | Five treating therapists (nominal code); caseloads of 41, 40, 27, 20 and 6 patients; not recorded for 7 patients |

At the time of referral, based on ICD-10 coding, patients presented with a diagnosis of mild cognitive impairment (87/141, 61.7%), dementia (41/141, 29.1%) or neither (13/141, 9.2%). Other diagnoses noted upon referral included sequelae of a cerebrovascular accident (CVA) (8/141, 5.7%), traumatic brain injury (TBI) or post-concussive injury (9/141, 6.4%), Parkinson’s disease (11/141, 7.8%), multiple sclerosis (MS) (3/141, 2.1%) or other diagnoses (12/141, 8.5%), which were not mutually exclusive. Patients included in the cohort analysis were assessed at the start of therapy and had mild-to-moderate cognitive impairment (Montreal Cognitive Assessment score of 21.1±4.0; Global Deterioration Scale (GDS) of 3.1±0.5).

Patients were highly compliant in completing the recommended three automated calls of cognitive exercises per week through Moneta’s digital therapy platform, with a mean of 2.7±0.8 exercise calls completed per week. Duration of treatment was a mean of 9.6±4.4 weeks in the program (range of 4 to 24 weeks). Thirty-one percent of patients had a care partner such as a family member participate with them in the program.

### Change in Pre-post Outcome Measures

Pre-post comparisons of outcome measures revealed significant improvements (Δ) from initial evaluation (PRE) to discharge (POST) for patients across all outcomes, with medium to very large effect sizes based on Cohen’s *d* noted for all measures, as shown in Figure 1 and Table 3. For all outcome measures evaluated, a majority of patients saw an improvement over the course of treatment. Average observed changes in FCM outcome measures exceed the benchmarks extracted from the ASHA NOMS registry for a roughly age-matched population being treated for cognitive communication disorders. The proportion of patients who met or exceeded the national benchmark was 53.7%, 49.4% and 57.4% for FCM-Cognition, FCM-Expressive Language and FCM-Language Comprehension, respectively.

**Table 3.** Change in outcome measures over the course of treatment and comparison to national benchmarks. N = sample size; Δ = delta or change in outcome measure; SD = standard deviation; CI = 95% confidence interval for the mean change (t-distribution); p from paired t-test (Neuro-QoL) or Wilcoxon signed-rank test (FCMs); % Improving = proportion of patients with Δ > 0; ASHA Benchmark = benchmark data corresponding to American Speech-Language-Hearing Association National Outcomes Measurement System (ASHA NOMS) database average outcomes extracted on July 9, 2025.

| Outcome Measure | N | Study $\Delta$<br>(mean±SD) | 95% CI | Effect Size<br>(Cohen's d) | % Improving | p | ASHA NOMS Benchmark $\Delta$ |
| --- | --- | --- | --- | --- | --- | --- | --- |
| Neuro-QoL | 116 | 4.0±5.6 | [3.0, 5.1] | 0.724 | 70.7% | <0.001 | - |
| FCM-Cognition | 134 | 18.2±18.6 | [15.1, 21.4] | 0.981 | 80.6% | <0.001 | 13.0 |
| FCM-Language Comprehension | 54 | 18.8±14.4 | [14.8, 22.7] | 1.303 | 94.4% | <0.001 | 14.7 |
| FCM-Expressive Language | 83 | 13.8±14.1 | [10.7, 16.9] | 0.979 | 85.5% | <0.001 | 12.2 |

**Figure 1.**
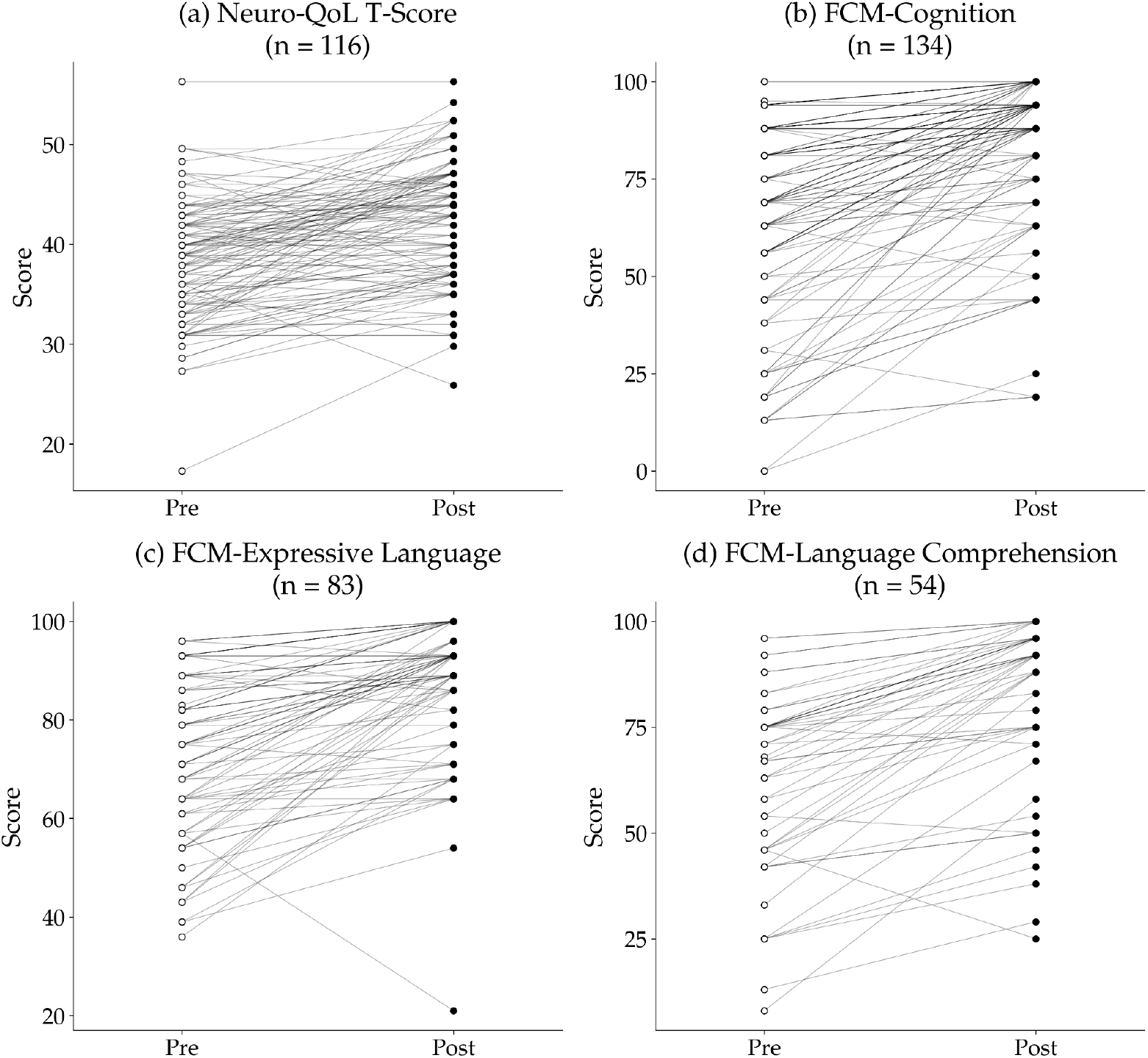
Individual pre and post values in outcome measures over the course of virtual CR treatment. Values shown were collected at the time of intake (PRE) and at the time of discharge (POST) for cohort population where PRE/POST data was available for the (a) Neuro-QoL T-Score (*N*=116), (b) FCM-Cognition (*N*=134), (c) FCM-Expressive Language (*N*=83), and (d) FCM-Language Comprehension (*N*=54). *Neuro-QoL = Quality of Life in Neurological Disorders; FCM = Functional Communication Measure*.

### Modeling Change in Outcome Measures

Models with meaningful predictive power were identified for FCM-Cognition and FCM-Expressive Language, a statistically significant but weak model (CV *R*^*2*^ = 0.064) for the Neuro-QoL, and no predictive model for FCM-Language Comprehension, as shown in Table 4. Best performing predictive models for changes in the Neuro-QoL, FCM-Cognition and FCM-Expressive Language showed statistical significance (*p*<0.05). Predictability varied substantially across the four targets, and sample sizes ranged from 48 to 128 patients.

**Table 4.**
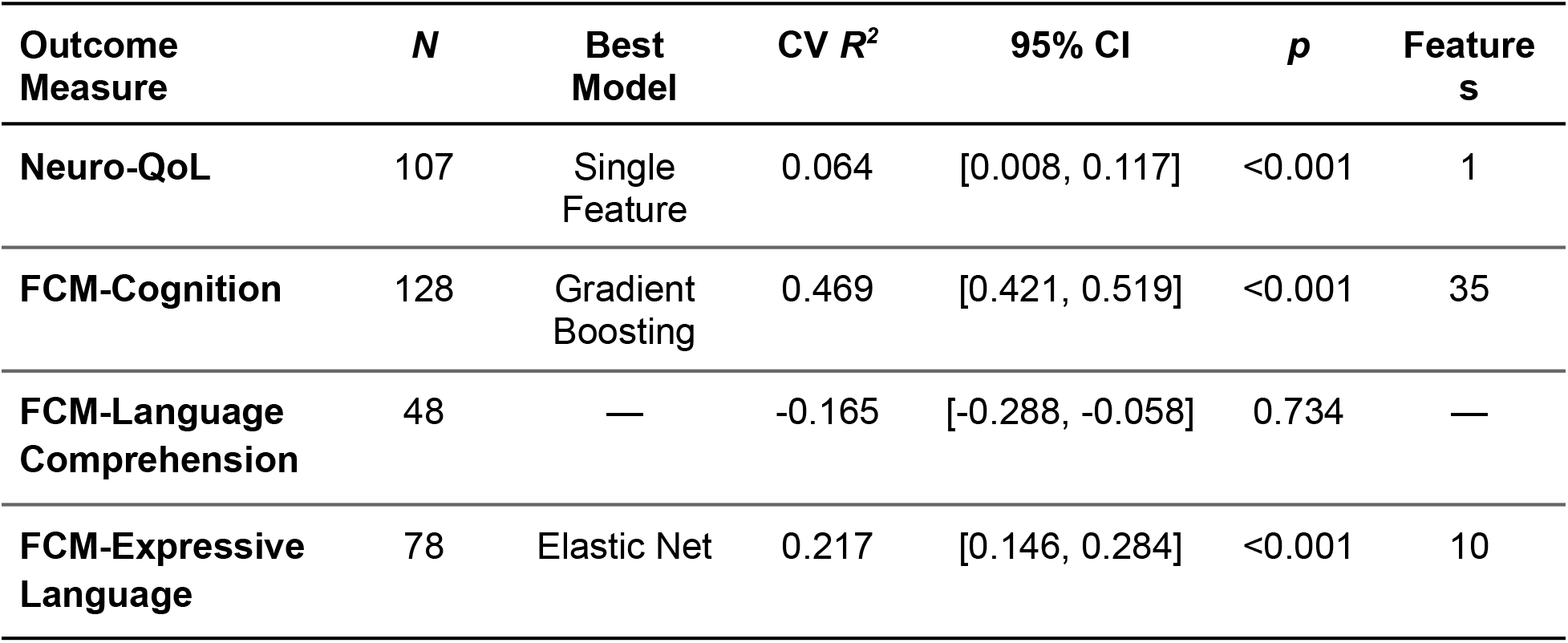
Summary of models that best predict change in each clinical outcome measure. Best model was selected by cross-validated R^2^; negative values indicate no predictive signal (performance worse than predicting the sample mean). N = number of patients included; Best Model = model type; CV R^2^ = mean cross-validated coefficient of determination; CI = 95% bootstrap confidence interval; p = one-tailed permutation p-value (1,000 permutations); Features = number of predictors used by the model (all 35 candidates for Gradient Boosting, which performs no feature selection; the retained subset for linear models). For FCM-Language Comprehension no model is selected; the CV R^2^ shown is the best of the six candidates.

| Outcome Measure | N | Best Model | CV $R^2$ | 95% CI | p | Features |
| --- | --- | --- | --- | --- | --- | --- |
| Neuro-QoL | 107 | Single Feature | 0.064 | [0.008, 0.117] | <0.001 | 1 |
| FCM-Cognition | 128 | Gradient Boosting | 0.469 | [0.421, 0.519] | <0.001 | 35 |
| FCM-Language Comprehension | 48 | — | -0.165 | [-0.288, -0.058] | 0.734 | — |
| FCM-Expressive Language | 78 | Elastic Net | 0.217 | [0.146, 0.284] | <0.001 | 10 |

### Predictors for Change in Overall Cognitive Abilities

The outcome measure FCM-Cognition is relevant to the full patient population as it assesses overall cognitive-related abilities in daily life. To further investigate predictor weighting, different models were compared for FCM-Cognition (see Supplemental Data - Table A1). All models were statistically significant, with the top performing model explaining the most variance in the sample being Gradient Boosting with 35 features (CV *R*^*2*^ = 0.469, 95% CI = [0.421, 0.519], *p* <0.001).

### Predictors for Change in Quality of Life

Models to predict changes in subject quality of life using the Neuro-QoL over the course of virtual CR showed significance (*p*<0.01) in five of six models (see Supplemental Data - Table A2). A univariate model using the baseline value of the Neuro-QoL T-Score outperformed all multi-feature models (CV *R*^*2*^ = 0.064, 95% CI = [0.008, 0.117], *p* <0.001). The addition of demographic or treatment-related features produced no improvement in model performance. This was the only target for which ordinary least squares assumptions were satisfied; among the linear models, AICc favored a three-feature Bayesian Ridge model, although its CV *R*^*2*^ (0.016) remained below the single-feature baseline, reflecting the parsimony-versus-fit trade-off when the underlying signal is weak.

### Predictors for Change in Language Comprehension

Models for changes in FCM-Language Comprehension showed no predictive value (*N* = 48; see Supplemental Data - Table A3). All models evaluated produced a negative *R*^*2*^ (best performing was Elastic Net: CV *R*^*2*^ = -0.165; 95% CI = [-0.288, -0.058]; *p* = 0.734). No multi-predictor model performed better than chance on permutation testing. A negative *R*^*2*^ indicates that the models predicted individual change less accurately than simply assigning the mean change to every patient and failed to generalize to the held-out dataset.

### Predictors for Change in Expressive Language

Permutation testing for FCM-Expressive Language was significant for all models (*p*<0.05), indicating detectable signal (*N* = 78, see Supplemental Data - Table A4). Elastic Net with 10 features was the best performing model (CV *R*^*2*^ = 0.217; 95% CI = [0.146, 0.284]; *p* <0.001); the fold-to-fold standard deviation (±0.240) was comparable to the mean estimate. Individual-level predictions remained imprecise. Within the Elastic Net model, the top three dominant predictive features of an increase in FCM-Expressive Language were baseline cognitive function (negatively weighted), treating therapist (positively weighted) and average weekly automated therapy calls (positively weighted). Notably, baseline cognitive function (FCM-Cognition (PRE)) carried the largest weight in the Elastic Net equation, whereas the strongest single predictor in the univariate baseline was the indicator for one treating therapist (CV *R*^*2*^ = 0.104). The second therapist indicator retained in the equation corresponds to a single patient and should not be interpreted.

## Discussion

Access to practical and personalized therapy is a critical component to helping older adults with cognitive impairment to remain independent in daily life. Support for verbal communication is already a core component of SLP practice for dementia due to a variety of underlying causes, addressing earlier stage deficits such as word finding, language comprehension and memory retrieval leveraging reserved capabilities. This study examined if a new model of virtual CR – designed to build on established SLP practice through a more accessible, telephone-based delivery system – produced clinically meaningful improvement in key outcome measures related to dementia progression and care. The key finding of significant and meaningful improvement in clinical outcomes supports expansion of real-world implementation of virtual CR, particularly when in-person care is limited. Furthermore, we explored if the change in the clinically meaningful outcomes could be predicted by either uni- or multi-variate modeling.

### Virtual CR is Associated With Improved Cognitive Function and Quality of Life

The core purpose of CR led by SLPs is to help individuals stay independent by compensating for cognitive deficits, maintaining effective communication and social engagement throughout the lifespan^18^. A key purpose of CR is also to maintain quality of life through sustaining participation and engagement in the patient’s preferred activities. Given the personalized delivery of CR focused on achieving these goals with the patient, and evidence from in-person models of CR, we can reasonably expect functional abilities in daily life and quality of life to improve over the course of therapy^3^. Indeed, through this retrospective analysis of real-world patient data, we observed significant improvement in therapist-assessed functional communication measures and patient-reported quality of life (Neuro-QoL) across the treatment period, i.e. intake to discharge. These findings demonstrate that clinically meaningful and practical gains can be achieved through a remotely delivered telehealth approach that includes sessions with both therapists and automated exercises.

While, to our knowledge, this is the first study to evaluate the impact of *telephone-based* cognitive rehabilitation therapy, these results are consistent with other studies evaluating the impact of *in-person* cognitive rehabilitation therapy on functional abilities for individuals with MCI or early dementia. Therapist-led cognitive interventions have been associated with improvements in generalizable outcomes such as activities of daily living, mood, and quality of life compared to controls^19^. A large-scale randomized trial showed significantly lower functional disability and a six-month delay in institutionalization for individualized, in-person CR for early-stage dementia at 24 months^6^. A review of the effects of cognitive remediation interventions, which includes CR as well as cognitive training and stimulation, provided evidence of improved instrumental activities of daily living for individualized therapy but showed insufficient evidence of a lasting effect^20^. These results underscore the need for longer-horizon outcome data to establish whether gains are maintained for a highly accessible, telephone-based CR model.

### Changes in Outcomes Are Comparable to National Benchmarks for In-Person Treatment

The relevance of these gains in clinical outcomes is reinforced by comparison with national benchmarks established by data contributed by US-based SLPs treating patients with cognitive communication disorders inclusive of MCI and dementia. In this study, average improvements on all three therapist-rated functional communication measures (FCMs) met or exceeded the corresponding ASHA NOMS benchmarks for outpatient, in-person treatment of cognitive-communication disorders. Although this comparison does not represent a matched control population, it provides evidence that telephone-based CR is a viable option for patient care demonstrating comparable benefit to in-person delivery within our patient population.

### Baseline Clinical Status Predicts Improvement in Cognitive Function and Quality of Life

An outstanding question for integrating CR into cognitive care pathways is which patients are likely to see benefit from the intervention. Here we take a modeling approach to identify what combinations of baseline characteristics provided the strongest association with changes in outcomes in a real-world population. While these results suggest that most patients stand to improve their cognitive function after CR, formulating a better understanding of the predictors of improvement may have value in identifying patients most likely to see meaningful gains.

Much of the predictive performance for FCM-Cognition is attributable to regression to the mean; the correlation between baseline FCM-Cognition (PRE) and change in the score over the course of treatment was strongly negative (r = -0.663). Patients with lower baseline cognitive function tended to show larger improvement irrespective of other factors. Residual diagnostics indicated pronounced heteroscedasticity, with residual variance approximately 17-fold greater among patients with low baseline FCM-Cognition (PRE) scores than among those near the upper bound of the scale. The increment in predictive performance attributable to other clinical and engagement features beyond baseline cognitive function (Δ*R*^*2*^ ≈ 0.10) is associated with influential features of treating therapist, age, total automated therapy calls and RIPA-G:2 raw score.

Baseline cognitive function (FCM-Cognition (PRE)) was a dominant predictor of change for FCM-Cognition and FCM-Expressive Language. The direction of this finding is consistent with prior research on the effect of a related cognitive intervention, cognitive stimulation, on people living with dementia, where the magnitude of improvement in cognitive function was negatively correlated with baseline cognitive function^21^. Although this result likely reflects a ceiling effect of the outcome measures, it may also indicate that patients with low baseline scores have greater room for measurable change. A lower level of cognitive function at baseline may offer more diversified avenues to successful intervention and improvement due to the highly personalized nature of therapist-led CR in addressing patient goals and adapting treatment strategy to ensure continued progress.

Efforts to predict an improvement in quality of life showed some insights but limited predictive value. The best performing model explained only around 6% of the variance, with the remaining variance attributable to measurement variability and factors not captured by our feature set. Although patient self-report of quality of life is valid in this population, corroboration by care partners could further improve data quality^22^. Nonetheless, the emergence of baseline quality of life as the primary predictor is consistent with other studies on predicting benefit from cognitive stimulation over eight weeks for individuals living with dementia in long-term care^21^. Interestingly, one controlled study that showed significant reduction in functional disability for patients receiving CR did not show a significant impact on change in quality-of-life at 3 or 24 months based on a traditional delivery model^6^. Although the drivers of change in quality-of-life remain poorly characterized in this population, the observation of a significant improvement is itself noteworthy and warrants further study.

Factors related to participant engagement with cognitive exercises delivered by a voice agent between therapist visits also surfaced as relevant to clinical benefit, underscoring the role of patient self-agency and the role of technology in scaling virtual CR. Overall treatment exposure in virtual CR as represented by the total number of completed automated cognitive exercise calls was a positive predictor of improvement in cognitive function, while intensity of treatment as represented by the average weekly automated calls was a predictor of gains in expressive language. Although the formats are materially different, this association is consistent with evidence of a dose–response relationship between computerized cognitive training and cognitive improvement, in which greater exposure to repetitive practice supports better outcomes, including for both MCI and mild dementia^23^. In healthy adults, higher engagement with training tools for strategy-based learning, coaching and brain-health habits was reported to be associated with the greatest gains in long term cognitive function over three years regardless of demographic factors^9^. These results suggest that greater overall exposure to and intensity of care enabled by technology may support increased generalization to daily life.

The emergence of treating-therapist identity as a predictor may reflect inter-clinician variation in outcome rating and program delivery given the highly individualized nature of the treatment and the subjectivity involved in clinical interpretations. Individual clinicians can account for a meaningful share of outcome variance within the broader therapy domain^24^. While inter-rater reliability of SLPs using the FCMs has been shown to be generally high, evidence points to some variability in how individual clinicians determine FCM ratings^25^. However, this study included a small number of participating therapists (five), limiting the interpretability of this result. In addition, one of the two therapist indicators retained in the expressive-language model represents a single patient and carries no interpretable weight.

### Limitations and Future Research

Although promising, we acknowledge several limitations to this study and areas of future investigation. The personalization of the CR format, consistent with standard practice by SLPs, introduces heterogeneity to the delivery model and treatment exposure, including program duration, cognitive exercise composition, live session format and patient engagement in automated exercises. We considered certain factors in our predictive modeling to understand their relative significance; number of automated therapy calls completed and treating-therapist identity both surfaced as meaningful predictors of cognitive functional improvement, with a greater number of completed calls being positively associated with improvement. A further consequence of using a single feature set across targets is that the models for FCM-Expressive Language and FCM-Language Comprehension were fit without their own baseline scores, which were too sparsely recorded to include without reducing every target to 30 or fewer patients (see Outcome Predictors). Given the strength of baseline effects for the other two outcomes, the predictors identified for expressive language should be interpreted with this omission in mind, and a re-analysis including the language baselines is warranted when a larger dataset is available.

Secondary diagnoses outside of MCI and dementia, such as Parkinson’s disease and traumatic brain injury, may be important factors in predicting clinical benefit from virtual CR. For example, a diagnosis of Parkinson’s disease was positively predictive of an improvement in expressive language in the best performing model. However, we caution against the generalizability of this finding as the dataset studied did not include sufficient samples within diagnostic subgroups to truly assess differences in clinical outcomes based on the diagnosis of comorbidities and warrants further analysis with a more representative and larger dataset.

Although these results provide a strong indication as to the overall benefit of virtual CR delivered by telephone in a real-world population of older adults, the study design did not include a direct comparison to traditional in-person CR, and therefore we are unable to determine equivalency or superiority. Finally, this retrospective analysis looked at pre-post outcome measures over the course of therapy but did not assess the long-term, post-treatment effects. Additional outcomes data at longer time horizons of up to two years and in combination with a maintenance program would test the sustainability of benefit.

## Conclusion

In summary, this real-world analysis provides initial validation of a telephone-based virtual CR model that pairs SLP-led therapy with automated exercises delivered by a voice agent, demonstrating significant functional and quality-of-life improvements, with functional gains that met or exceeded national in-person benchmarks. While the predictive model findings offer early guidance for identifying patients best suited for referral and for optimizing program execution, most patients evaluated in this analysis benefited from the virtual CR program, and roughly half met or exceeded the in-person benchmarks for clinical outcomes. The model’s low technological requirements alongside validated results position it well for extending this novel delivery model for virtual CR into rural and underserved communities.

## Data Availability

All data produced in the present study are available upon reasonable request to the authors.

## Author Contributions

Conceptualization: JF, MB, CS, JN; Methodology: JF, MB, CS, JN, ER; Data Analysis: ER, MB; Writing – Original Draft: JF, ER; Writing – Review and Editing: JF, JN, CS, MB, ER; Supervision: JF, MB; Project Administration: JF, CS, MB.

## Data Availability

The data presented in this study are available on request from the corresponding author. The data are not publicly available due to confidentiality restrictions.

## Acknowledgments

During the preparation of this manuscript, the authors used Anthropic Claude, Version Opus 4.8 (Anthropic) for the purposes of generating Figure 1 and text editing and grammar. The authors have reviewed and edited the output and take full responsibility for the content of this publication.

