## Supplemental Data Tables and Figure for "Real-World Outcomes of a Telephone-Based Virtual Cognitive Rehabilitation Therapy Program: A Retrospective Cohort Analysis"

#### Section A1. Comparison of Models for Predicting Change in Cognitive Function

Table A1 provides a comparison of models evaluated to predict change in cognitive function equal to FCM-Cognition (POST) minus FCM-Cognition (PRE) (*N* = 128). The Gradient Boosting model was the best performing model with the top five features (weighted importance) of FCM-Cognition (PRE) (0.4975); Treating Therapist Code – Therapist 3 indicator (0.1382); Age (0.1177); Total Automated Therapy Calls (0.0558); RIPA-G:2 score (0.0306).

**Table A1.** Comparison of models for predicting change in cognitive function during treatment. *CV = cross validation; SD = standard deviation; RMSE = root mean square error; CI = 95% bootstrap confidence interval;* p *is one-tailed permutation test value; Features = number of predictors used out of 35 candidates; CV $R^2$ and SD are across 5 × 10 repeated CV folds.*

| Model | CV $R^2$ | ±SD | RMSE | 95% CI | *p* | Features |
|---|---|---|---|---|---|---|
| **Gradient Boosting** | 0.469 | 0.171 | 12.911 | [0.421, 0.519] | <0.001 | 35 |
| **Lasso** | 0.462 | 0.168 | 13.002 | [0.411, 0.507] | <0.001 | 10 |
| **Elastic Net** | 0.443 | 0.142 | 13.330 | [0.403, 0.482] | <0.001 | 3 |
| **Bayesian Ridge** | 0.433 | 0.197 | 13.289 | [0.374, 0.485] | <0.001 | 3 |
| **Ridge** | 0.415 | 0.172 | 13.614 | [0.366, 0.462] | <0.001 | 5 |
| **Single Feature** | 0.370 | 0.167 | 14.188 | [0.323, 0.416] | <0.001 | 1 |

#### Section A2. Comparison of Models for Predicting Change in Quality of Life

Table A2 provides a comparison of models developed to predict change in quality of life equal to Neuro-QoL T-Score (POST) minus Neuro-QoL T-Score (PRE) (rounded to whole T-score points, as recorded in the dataset) (*N* = 107). The Single Feature model was the best performing model represented by Equation 1 (with equation coefficients on standardized predictors):

$$y = 4.159 - 2.352 \times \text{Neuro-QoL T-Score (PRE)} \qquad (1)$$

**Table A2.** Comparison of models for predicting change in quality of life during treatment. *CV = cross validation; SD = standard deviation; RMSE = root mean square error; CI = 95% bootstrap confidence interval;* p *is one-tailed permutation test value; Features = number of predictors used out of 35 candidates; CV $R^2$ and SD are across 5 × 10 repeated CV folds.*

| Model | CV $R^2$ | ±SD | RMSE | 95% CI | *p* | Features |
|---|---|---|---|---|---|---|
| **Single Feature** | 0.064 | 0.205 | 5.015 | [0.008, 0.117] | <0.001 | 1 |
| **Lasso** | 0.037 | 0.137 | 5.126 | [-0.001, 0.074] | <0.001 | 5 |
| **Elastic Net** | 0.027 | 0.139 | 5.151 | [-0.011, 0.064] | <0.001 | 3 |
| **Ridge** | 0.017 | 0.142 | 5.170 | [-0.024, 0.055] | <0.001 | 3 |
| **Bayesian Ridge** | 0.016 | 0.210 | 5.151 | [-0.042, 0.070] | 0.002 | 3 |
| **Gradient Boosting** | -0.349 | 0.430 | 5.928 | [-0.476, -0.231] | 0.065 | 35 |

**Section A3. Comparison of Models for Predicting Change in Language Comprehension**

Table A3 provides a comparison of models developed to predict change in language comprehension equal to FCM-Language Comprehension (POST) minus FCM-Language Comprehension (PRE) ($N$ = 48). No multi-predictor model performed better than chance on permutation testing.

**Table A3.** Comparison of models for predicting change in language comprehension during treatment. *CV = cross validation; SD = standard deviation; RMSE = root mean square error; CI = 95% bootstrap confidence interval;* p *is one-tailed permutation test value; Features = number of predictors used out of 35 candidates; CV $R^2$ and SD are across 5 × 10 repeated CV folds.*

| Model | CV $R^2$ | ±SD | RMSE | 95% CI | *p* | Features |
|---|---|---|---|---|---|---|
| **Elastic Net** | -0.165 | 0.417 | 14.461 | [-0.288, -0.058] | 0.734 | 7 |
| **Lasso** | -0.253 | 0.427 | 15.119 | [-0.378, -0.140] | 0.892 | 3 |
| **Gradient Boosting** | -0.274 | 0.773 | 14.604 | [-0.515, -0.076] | 0.231 | 35 |
| **Ridge** | -0.278 | 0.936 | 14.650 | [-0.564, -0.072] | 0.754 | 10 |
| **Bayesian Ridge** | -0.293 | 1.063 | 14.349 | [-0.647, -0.050] | 0.469 | 10 |
| **Single Feature** | -0.488 | 0.727 | 16.009 | [-0.693, -0.304] | 0.030 | 1 |

### Section A4. Comparison of Models for Predicting Change in Expressive Language

Table A4 provides a comparison of models developed to predict change in expressive language equal to FCM-Expressive Language (POST) minus FCM-Expressive Language (PRE) ($N = 78$). The Elastic Net model was the best performing model represented by Equation 2 (with equation coefficients on standardized predictors):

y = 13.590 + 2.567 × Average Weekly Automated Therapy Calls + 1.238 × MoCA Score − 4.573 × FCM-Cognition (PRE) + 4.092 × Treating Therapist-Therapist3 + 1.891 × Treating Therapist-Therapist5 + 1.620 × Sex − 1.250 × Diagnosis-Dementia + 1.797 × Diagnosis-TBI + 1.751 × Diagnosis-PD − 0.627 × Current Living Environment-At Home with Paid Support (2)

**Table A4.** Comparison of models for predicting change in expressive language during treatment. *CV = cross validation; SD = standard deviation; RMSE = root mean square error; CI = 95% bootstrap confidence interval;* p *is one-tailed permutation test value; Features = number of predictors used out of 35 candidates; CV $R^2$ and SD are across 5 × 10 repeated CV folds.*

| Model | CV $R^2$ | ±SD | RMSE | 95% CI | $p$ | Features |
|---|---|---|---|---|---|---|
| **Elastic Net** | 0.217 | 0.240 | 12.026 | [0.146, 0.284] | <0.001 | 10 |
| **Lasso** | 0.203 | 0.273 | 12.059 | [0.121, 0.277] | <0.001 | 10 |
| **Gradient Boosting** | 0.181 | 0.308 | 12.150 | [0.092, 0.267] | <0.001 | 35 |
| **Bayesian Ridge** | 0.148 | 0.375 | 12.253 | [0.041, 0.251] | <0.001 | 10 |
| **Single Feature** | 0.104 | 0.276 | 12.898 | [0.029, 0.184] | <0.001 | 1 |
| **Ridge** | 0.075 | 0.306 | 12.954 | [-0.017, 0.159] | <0.001 | 10 |

### Section A5. Correlation of Characteristics in Study Population

The clinical baseline measures of the MoCA, RIPA-G:2, and FCM-Cognition (PRE) showed moderate to strong positive inter-correlations, as shown in Figure A5. The GDS, which is an overall measure of the progression of cognitive decline and a higher score indicates greater severity, was negatively correlated with the MoCA, RIPA-G:2 and baseline FCM-Cognition (PRE), where a higher score generally indicates better cognitive functioning. A higher baseline patient-reported quality of life, Neuro-QoL T-Score (PRE), was positively correlated with higher baseline cognitive function (FCM-Cognition) and inversely correlated with the severity of cognitive decline (GDS), showing expected convergence between clinician and patient-reported functional measures.

Demographic factors also showed correlations with clinical factors. More advanced age was associated with a higher baseline quality-of-life and hearing impairment in the cohort. Educational level was inversely associated with the overall severity of cognitive decline (GDS) and positively associated with measures of cognition (MoCA and RIPA-G:2). Patient diagnoses, age and hearing impairment were not strongly correlated with engagement in the program, nor were clinical baseline measures of cognition.

The categorization of patients into an MCI or dementia diagnosis showed moderate association with clinical measures of cognition at baseline. A dementia diagnosis was negatively correlated with baseline cognitive function as measured by the MoCA, RIPA-G:2 and the baseline clinical measures of FCM-Cognition (PRE) and Neuro-QoL T-Score (PRE). A diagnosis of MCI (exclusive of a dementia diagnosis) showed the opposite pattern, with positive correlations with these measures and a negative correlation with the GDS.

**Figure A5. Predictor correlation matrix.** *Pearson's correlations between all predictors including population demographics, clinical features and baseline outcome measures (N = 131). Red indicates positive correlation whereas blue indicates a negative correlation. Wks = weeks; Wkly = weekly; Avg = average; auto = automated; MoCA = Montreal Cognitive Assessment; RIPA-G:2 = Ross Information Processing Assessment - Geriatric, Second Edition; GDS = Global Deterioration Scale; Dx = diagnosis; MCI = mild cognitive impairment; MS = multiple sclerosis; TBI = traumatic brain injury; CVA = cerebrovascular accident/stroke; Imp. = impairment.*

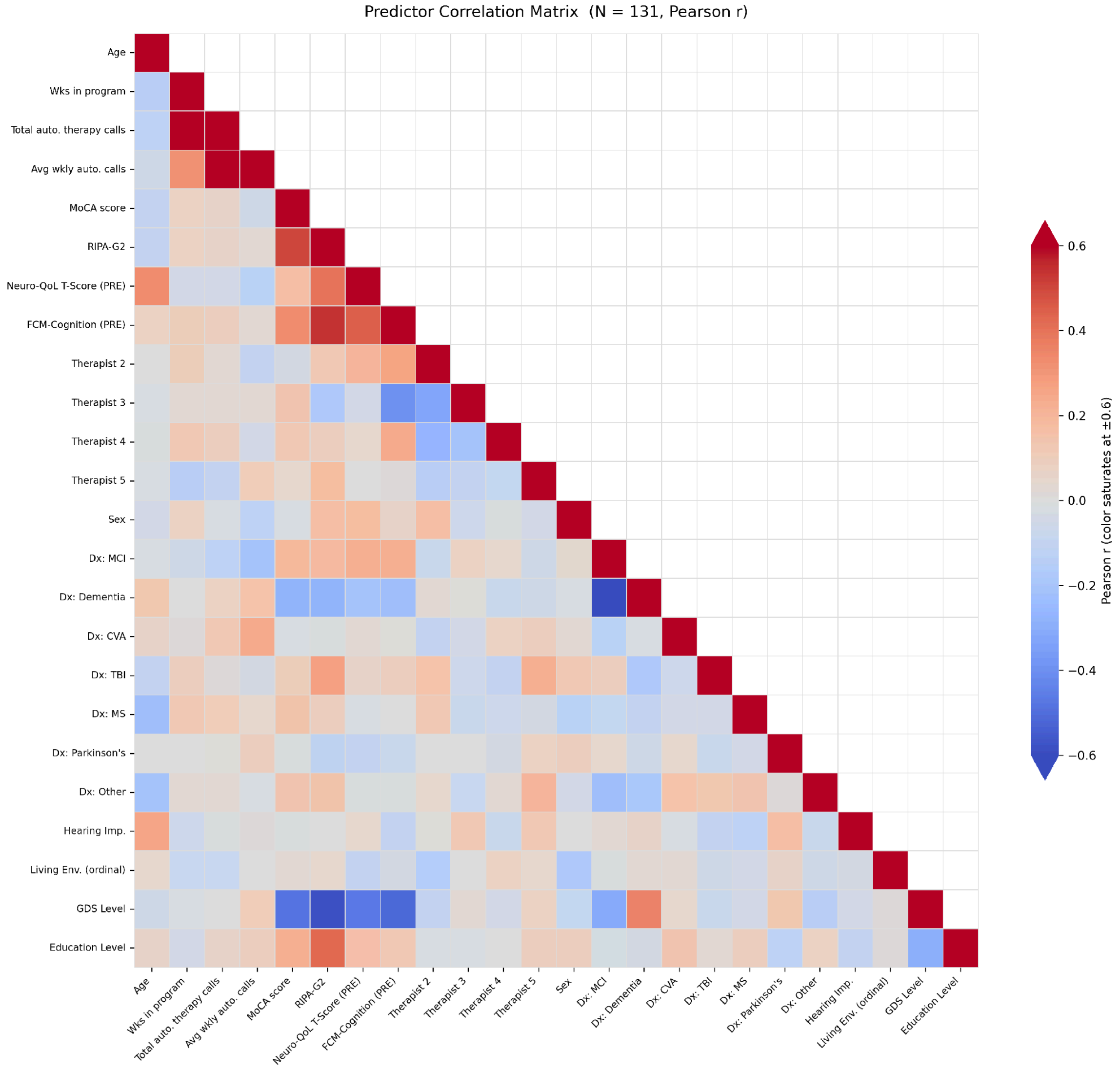
